# Design and implementation of Electronic Health Record Computerised Decision Support System (CDSS) trials: A Systematic Review

**DOI:** 10.1101/2024.10.25.24316128

**Authors:** Yang Chen, Matthew G. Wilson, Myura Nagendran, Didier Collard, Jayathri Wijayarathne, Matiwalakumbura Dilan, Ravi Wickramaratne, Karan Dahele, Jack Samways, Yogini Jani, Anoop Shah, Tom Lumbers, Steve Harris, Folkert W Asselbergs

## Abstract

**Objectives:** Conduct a systematic review of the existing evidence base pertaining to the conduct of randomised controlled trials of clinical decision support systems embedded within electronic health record systems. Further, to describe whether key features of trial design and implementation were consistently reported.

**Materials and Methods:** A systematic search of MEDLINE was conducted in April 2022. Three independent reviewers screened the search results. A 27-item checklist was used to extract data from the screened studies. A subgroup analysis was conducted to classify trials of clinical decision support systems based on whether they encouraged guideline adherence or represented new knowledge generating mechanisms.

**Results:** 5,213 records were retrieved. Following screening, 106 studies were included in the review. The majority of studies evaluated active alerts seeking to improve adherence to clinical guidelines rather than generate new knowledge. Few studies quantified the existing ecosystem of decision support at the study site, or explored phenomena like alert fatigue.

**Discussion:** This systematic review provides a detailed analysis of the characteristics of trials evaluating clinical decision support systems. It highlights significant under-reporting of key factors which may affect the reproducibility and generalisability of trial results - particularly with respect to measurement of alert fatigue, description of the underlying digital ecosystem and additional co-interventions used within trials.

**Conclusion:** As clinical workflows undergo digital transformation, randomised controlled trials of clinical decision support systems require greater standardisation, in both conduct and reporting. This represents an area of expanding interest given the increasing use of artificial intelligence-enabled decision support.

**STRENGTHS & LIMITATIONS:**

- This study presents the results of an updated systematic review of studies evaluating the effectiveness clinical decision support systems.
- It used a comprehensive checklist to extract detail pertaining to five information domains on trial quality and description.
- Studies were evaluated to determine whether the clinical decision support system was knowledge generating or designed to improve guideline adherence.
- The review was limited to randomised trials and excluded quasi-experimental and observational studies of clinical decision support systems.

## BACKGROUND & SIGNIFICANCE

Clinical decision-making will increasingly incorporate support from artificial intelligence (AI) algorithms.(1) Most AI algorithms will assist clinician decision-making rather than act as autonomous agents(2) and will therefore require integration into clinical pathways through computerised decision support systems (CDSS). While many of the AI-specific challenges to evaluating AI-CDSS have been highlighted through reporting guidelines such as DECIDE-AI (3), there remain significant uncertainties in determining the features of CDSS design and implementation that are AI-independent and which are fundamental to the evaluation and generalisability of trial results. This work seeks to address this important gap in the literature.

CDSS have been the subject of research and implementation efforts for decades(4), and can be categorised by whether they support existing best practice (guideline adherence) or seek to generate new knowledge. A 2020 meta-analysis which focused on CDSS aimed at improving guideline adherence reported an overall effect of 6% absolute improvement in patient care.(5) However, this assertion was based on heterogenous studies with significant variation in clinical context, outcomes, and a lack of detail in key elements of trial design and implementation that could limit the generalisability of the purported CDSS effect. In addition, given the narrow focus on guideline adherence trials, no observations were made regarding whether knowledge generating CDSS require different levels of consideration in terms of study design and implementation. Specifically, no comparisons were made between a simple guideline-based reminder to a complex decision support system seeking to address novel research questions.(6)

Our work aims to provide a contemporary synthesis of the salient features reported in existing randomised controlled trials (RCTs) of CDSS. We specifically examine the reporting of the types of CDSS, details of local implementation, and key elements of study design including models of consent and descriptions of co-interventions or quantification of alert burden where relevant. As CDSS become increasingly available, complex and intelligent(7), and their development and testing increasingly regulated, a greater understanding of the essential requirements for their design, development and delivery within RCTs is of critical importance.

## METHODS

The protocol for this systematic review was registered on PROSPERO (CRD42022327682) prior to execution of the search. This manuscript has been prepared according to the updated 2020 guidelines issued by the PRISMA group and the SWiM extension.(8, 9) A checklist is available in the Supplemental appendices along with a list of protocol amendments.

### Eligibility criteria

Eligible studies included RCTs of a decision support intervention integrated within the electronic health record (EHR) clinical information system, routinely used by clinicians at the time of providing care to a patient (e.g., while entering an order or a clinical note). The comparator was standard of care.

### CDSS intervention

We only included studies where at least one of either input or output functions for the CDSS was built into the EHR used for clinical care. Applications that collected data separate to the EHR and presented these separately were excluded. Specialised diagnostic decision support systems e.g., in medical imaging and administrative systems such as billing or coding support were excluded. CDSS that had an indirect effect on clinical care e.g., improving clinician documentation, were also excluded. (10)

### Study identification, selection and data extraction

We elected to use the comprehensive search strategy by Kwan et al, and updated the time filter to run through from January 2010 through to April 2022.(5) The detailed search strategy is given in Appendix 1. After removal of duplicates, independent reviewers (DC, YC, MW) screened the titles and abstracts of the search results. The full texts of the remaining results were screened by two independent reviewers with arbitration by a third author if necessary (YC). Data from eligible studies was extracted from study reports independently by reviewers (YC, MW, JW, MD, RW, KD) with each study being assessed by two independent reviewers. Two papers from each scorer were selected at random and audited independently by a separate author (YC). If auditing revealed significant discrepancies, a re-evaluation of the original scoring was triggered. Zotero was used for reference management.

### Study reporting

For eligible studies, we report a modified 27-item list of data elements from our initial 23-item list outlined in the protocol. These were grouped into the following five domains reflecting our study aim: (i) design features and safety, (ii) decision context, (iii) study design and implementation, (iv) relevant human factors, (v) study outcomes and reporting. The final list of items chosen were based wherever possible on relevant reporting guidelines for clinical trials, computerised interventions and EHR use and a detailed breakdown is available in the supplement. (11, 12) (13) For each item, we present the number of studies which satisfy the conditions of that data element, rather than using a narrower metric such as adherence.

### CDSS and item classification

We defined CDSS interventions according to whether they supported adherence to known best practice (GDT - Guideline directed therapy) or whether they aimed to generate new knowledge insights (KG – knowledge generating). Examples of each category are available in Table 1. For certain items, further elaboration is provided in the supplemental materials (Appendix 3). For example, the classification of ‘active’ versus ‘passive’ alert was determined by whether the alert was displayed and interrupted users without requiring any action other than the triggering condition (active) or if there was a requirement to click on a separate element of the user interface (passive, non-interruptive). Active alerts were further divided into either hard stop (the user had to interact with the alert to proceed) or soft stop (the alert could be dismissed easily).

**Table 1.** Classification of decision support into those that aid guideline directed therapy (GDT) and those that are knowledge generating (KG).

| Guideline directed therapy (GDT) |  | Knowledge generation (KG) |  |
| --- | --- | --- | --- |
| <u>GDT- Simple</u> | <u>GDT - Novel</u> | <u>KG - Simple</u> | <u>KG - Novel</u> |
| Simple alert or recommendation e.g., prescribe anticoagulation for patients with a clear indication | New information given to clinician e.g., calculation of ChadsVasc to aid guideline directed therapy | Test novel hypothesis in areas of clinical practice variation. CDSS used to recommend or nudge clinical behaviour and evaluate comparative effectiveness of existing therapies e.g., oral fluid restriction in acute heart failure vs. free fluids | Test novel hypothesis and present new information e.g., prognostic data for HF displayed to clinical team, and measure clinical decisions and patient outcomes |
| PROMPT-HF (20) | ALERT-AF(29) | THIRST Alert (25) | REVEAL-HF (6) |

### Data Analysis

The results were grouped according to whether the CDSS was GDT or KG and a descriptive analysis of each trial and group summary was conducted according to our chosen reporting metrics. No risk of bias assessment, assessment for publication bias or quantitative synthesis was conducted.

## RESULTS

A total of 5,213 records were retrieved by electronic searches, last updated on April 9, 2022 (**Figure 1**). After removal of duplicates and irrelevant records, 398 full text articles were assessed of which 344 were excluded. A final list of 54 papers from the search was added to 52 papers which met the eligibility criteria from the original list of papers identified in a previous published systematic review.(5)

**Figure 1.**
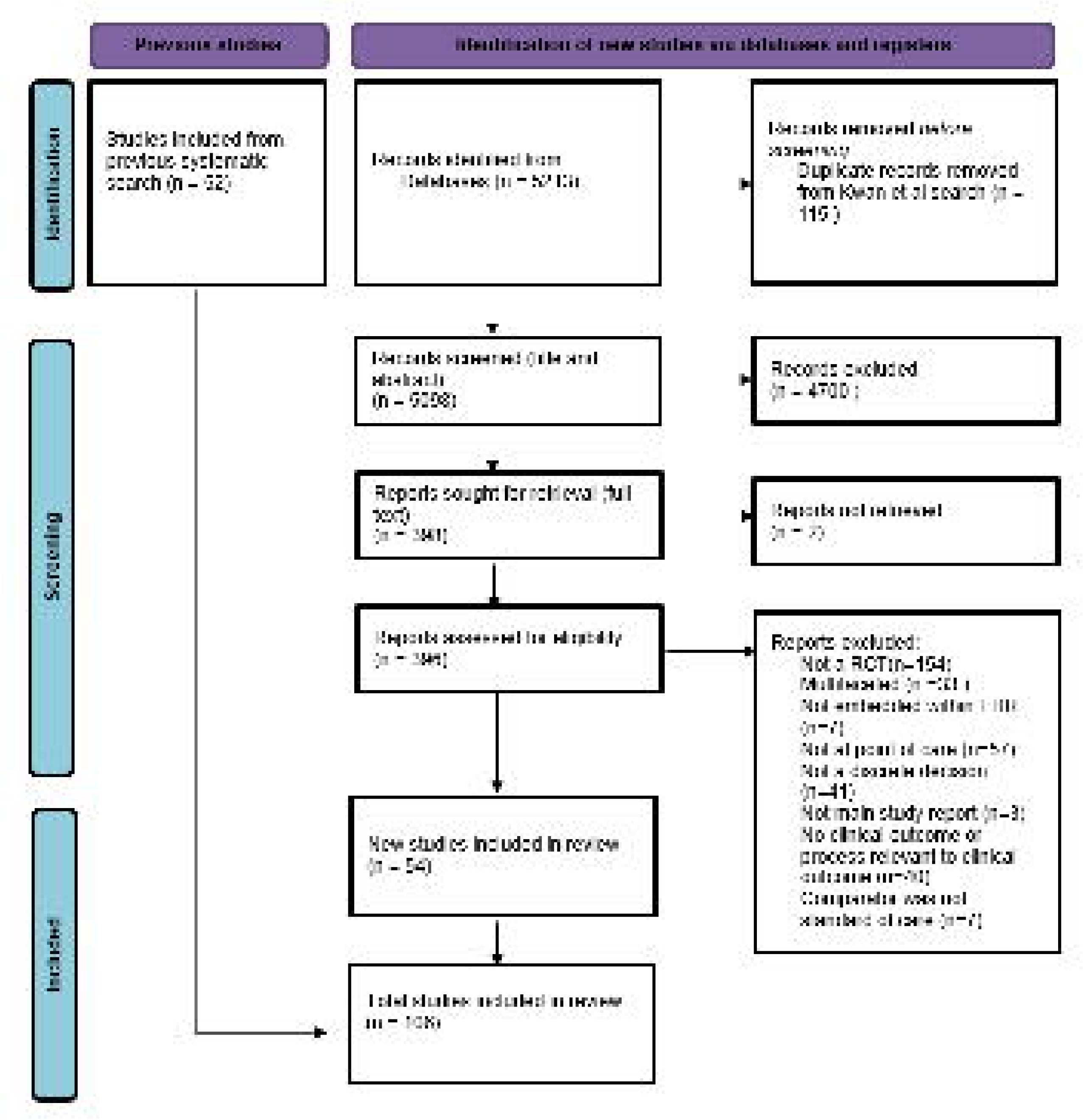
Study selection

The general characteristics of the 106 RCTs are summarised in **Table 2**. The overall clarity of reporting across the five domains varied significantly. For example within the domain of Design features and Safety, 3% of studies reported whether there was any ongoing monitoring of CDSS performance while 91% reported details about how the CDSS that was fully embedded into the EHR) (**Table 3**). 96 out of 106 RCTs (86%) used CDSS alerts and of these, 80 employed an active design. Most trials focused their intervention on either medically trained clinicians alone (49%) or multiple professional groups including medically trained clinicians (45%).

**Table 2.** Baseline characteristics of studies.

| Domain | Number of studies (%) |
| --- | --- |
| CDSS Form |  |
| Alert | 96 (91) |
| Template management protocol | 5 (5) |
| Template report | 2 (2) |
| Template order set | 1 (1) |
| Change in default order of medications presented | 1 (1) |
| Forcing function (removal of choices) | 1 (1) |
| CDSS Target |  |
| Physicians only | 52 (49) |
| Nurses or other professional group | 6 (6) |
| Multiple professional groups | 48 (45) |
| Type of decision supported |  |
| Guideline directed – simple | 64 (60) |
| Guideline directed – complex | 20 (19) |
| Knowledge generating – simple | 15 (14) |
| Knowledge generating – complex | 7 (7) |
| Trial Geography |  |
| USA | 71 (67) |
| Rest of world | 35 (33) |
| Cointerventions |  |
| None | 59 (56) |
| One | 34 (32) |
| Two or more | 13 (12) |
| Primary outcome |  |
| Process measure only | 60 (57) |
| Included Clinical measure | 46 (43) |

**Table 3.**
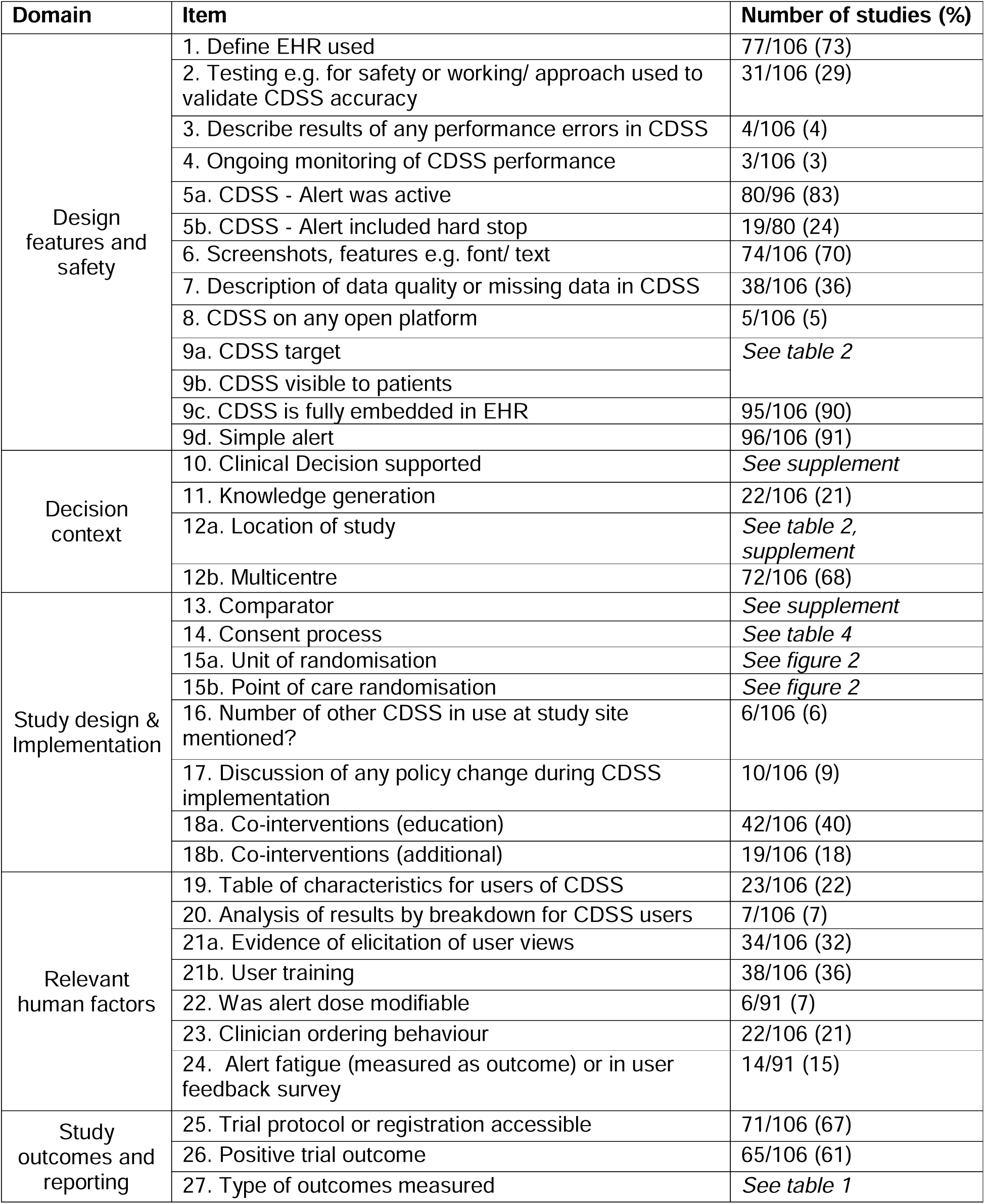
Reporting of included studies across domains of interest.

Only seven CDSS were additionally visible to the patient. 14 out of 96 alert-based studies (15%) included a measure of alert fatigue and only six studies referenced the presence or absence of other decision support tools within the EHR at the study site.

In terms of Decision context, 84 studies (79%) evaluated CDSS designed to improve adherence to guidelines. The majority of published RCTs were based in the US (71 of 106 studies). For study design and implementation, **Table 4** provides further detail about the different models of consent used in the studies. Of 50 studies with waiver of consent, 40 were conducted in the US (56% of US RCTs). There were significant differences in how randomisation was employed. 55 of 106 studies used a cluster randomization design while of the remaining 51 studies, only 16 used the EHR itself to conduct point of care randomisation (**Figure 2**). The use of co-interventions also varied significantly – 59 studies did not report any co-interventions such as staff education or regular feedback, 34 studies described the use of one co-intervention and 13 studies described two or more co-interventions.

**Table 4.** Different models of consent used.

|  | **Based in USA (%)** | **Rest of world (%)** | **Overall (%)** |
| --- | --- | --- | --- |
| **Waiver** | 40 (56) | 10 (26) | 50 (47) |
| **Clinical Lead** | 3 (4) | 4 (11) | 7 (7) |
| **Clinician** | 7 (6) | 8 (23) | 15 (14) |
| **Patient** | 2 (3) | 3 (9) | 5 (5) |
| **Patient & Clinician** | 5 (7) | 5 (14) | 10 (9) |
| **Not reported/ Unclear** | 14 (20) | 5 (14) | 19 (18) |

**Figure 2.**
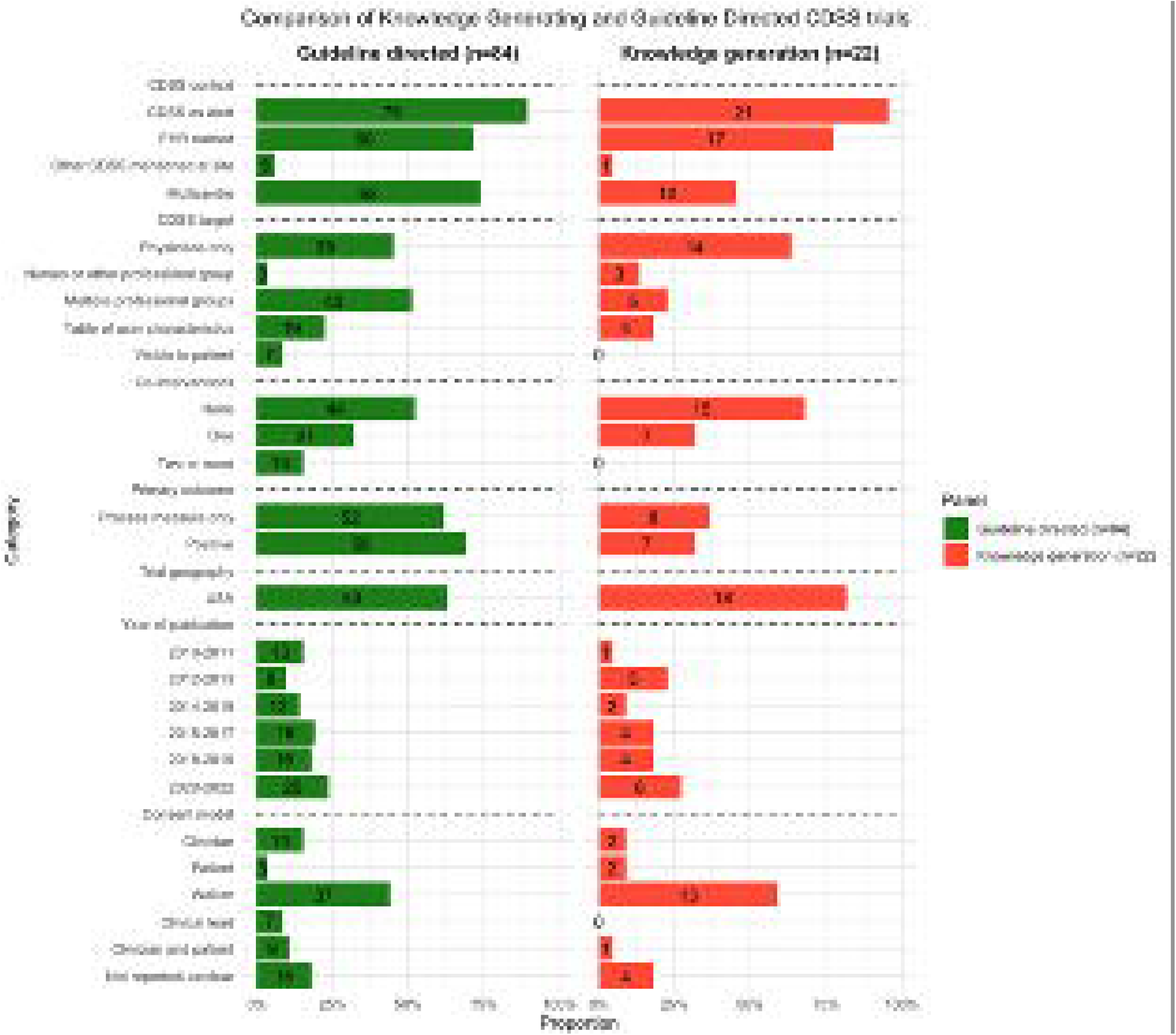
Randomisation procedure used in CDSS RCTs. Cluster randomisation was used in 55 studies (52%); patient in 24 studies (23%); clinician in 27 studies (25%). Of the 51 studies that randomised by either patient or clinician, only 16 (33%) reported that the EHR was used to conduct point of care randomisation

For human factors, only 23 of 106 studies (22%) presented a table of characteristics for different CDSS users and in terms of study outcomes, 65 of 106 studies reported a positive primary outcome though only 46 of 106 studies included a clinical measure.

**Figure 3** compares trial reporting according to whether the RCT was defined as knowledge generating (KG) or guideline directing (GDT). The proportion of studies that satisfied each domain element was similar between KG and GDT studies, with the following exceptions: (i) 64% of KG studies displayed the CDSS to physicians only compared to 45% of GDT studies, (ii) 68% of KG studies did not report using a co-intervention compared to 52% of GDT studies), (iii) (82% of KG studies were based in the US compared to 63% of GDT studies) and (iv) waiver of consent was used in 59% of KG studies compared to 39% of GDT studies.

**Figure 3.**
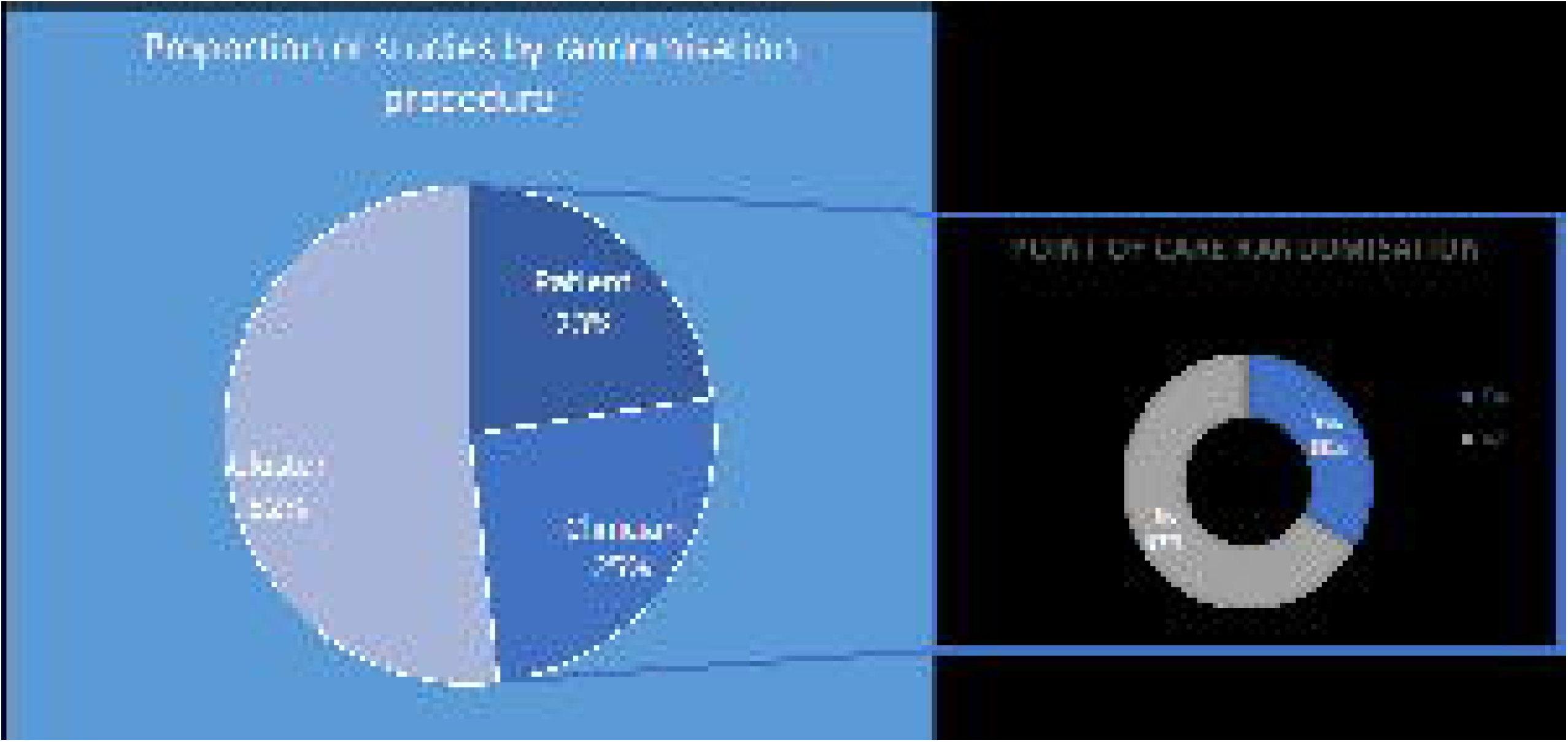
Selected characteristics of CDSS RCTs according to either knowledge generation or guideline directed therapy.

## DISCUSSION

We demonstrate in this systematic review that the majority of CDSS RCTs share the following features:

(i) They study the effect of an active alert embedded within the EHR and aim to improve compliance to guideline directed therapy.
(ii) They have common areas of underreporting which may affect their implementation at different study sites and limit reproducibility. Only 6 out of 106 studies referenced the existing decision support ecosystem at the study site in which the EHR-embedded CDSS was trialed.
(iii) There are few differences in the design and conduct between trials that aim to improve guideline directed therapy and those that seek to generate new knowledge insights. Exceptions include greater use of waiver of consent among KG trials, a focus on medically trained clinicians as their target and fewer co-interventions.

As the digital transformation of workflows spreads across healthcare, characterising and evaluating CDSS implementation when deployed as part of formal research studies, with or without AI, will become increasingly relevant to large multidisciplinary teams.

### Comparison to the literature

Whilst reporting standards or recommendations exist for areas such as trial reporting(13), decision aids(14), complex interventions(15) or the early-stage clinical evaluation of AI(3), no single checklist or framework addresses all of the factors relevant to determining how to effectively conduct research studies of CDSS and implement them within routine clinical workflows.

Our motivation was to highlight the residual gaps and uncertainty of the existing evidence base. Our analysis of CDSS design features, decision context, study design features such as randomisation and consent procedure, relevant human factors, and reported study outcomes were chosen based on their central role in determining the applicability of research findings across different healthcare systems.

As more trial evidence emerges to support the use of AI, particularly in medical imaging such as augmentation of clinical screening pathways (16) or as potential diagnostic agents, (17) the field of decision support will continue to grow and differentiate. Many digital tools in design or testing may therefore sit outside of traditional EHRs. The optimal integration of all tools within a changing clinical and digital workflow, will therefore not only require careful attention to design features and implementation as part of research studies or clinical practice, but also how structured data elements interact across platforms. Additional guidance from established frameworks such as SAFER: Sociotechnical Framework for Safety-Related Electronic Health Record Research Reporting(18) and CODE-EHR(11) should be incorporated into future benchmarks of study design and reporting.

### Limitations

In this review we have focused only on CDSS interventions designed for clinicians. Decision support in healthcare is an umbrella term and the subject of multiple previous systematic reviews, including Cochrane reviews of patient-facing decision aids.(19) There is lack of a standardised nomenclature to define the field of decision support, including digital or computerised decision support as applied to healthcare. While most trials in this review were of alerts, alternative forms of decision support are available, including changing defaults or forcing functions.(10) This heterogeneity limits the pooling of different CDSS designs and additionally restricts the ease of study identification, retrieval and publication as original research articles. Despite this, there is significant growth in the number of CDSS trials, particularly those using alerts. Since our search execution, prominent examples have included trials to assess whether CDSS can improve guideline adhering therapies in heart failure (PROMPT-HF)(20) or whether they can generate new knowledge insights (REVEAL-HF)(21). To balance comprehensiveness and feasibility, we elected to replicate the detailed strategy used by Kwan et al(5) and did not search additional trial registration platforms. We additionally chose not to perform a risk of bias assessment given this was already conducted in 52 of the RCTs from the previous review and our objectives were not to specifically assess the quality or validity of individual studies.(22)

### Practice implications

Decision support and AI offer the promise to transform healthcare and have accordingly received significant resource investment. However this is contradicted by the existing evidence base for foundational decision support systems where key operational details for interpreting their clinical effect and effective implementation are lacking. For any clinical workflow which aims to use a CDSS, determining the optimal way to introduce them as part of the ‘five rights’ (right information, right person, right form, right channel and right time) will require better reporting of future trials.(23)

For example, concerns about alert fatigue have been the subject of previous research(24), however in our search, only 15% of studies reported metrics that could indicate whether such a phenomenon was present. In addition, no study reported the granular detail of whether an alert was triggered in line with a clinician’s work (i.e., the required action or decision of the user matched the intended use of the CDSS) or whether the CDSS displayed distracting information while the clinician used the EHR for another purpose. Whether a CDSS design could ever satisfy the five rights for all users remains unclear. The current standard of reporting where fewer than one in ten trials present sub-analysis of CDSS effects by different user groups highlights the scale of challenge to present even basic information.

Rather than aiming to achieve the optimal timing for an ‘effective alert’, RCTs could introduce a preliminary step of harnessing active alerts to ‘nudge clinicians’ to consider whether to include a patient into a RCT in the first place.(25) The use of the term nudge raises separate issues about nomenclature and has been summarised elsewhere. (26)

Our results support the need to develop a greater understanding of the design and implementation of CDSS to explain why certain clusters of trials work as intended. Regardless of the complexity of design and code underpinning a given CDSS, and whether they include AI technology or not, all will rely on the complex interplay of factors which shape clinician behavioural (27), where rules of thumb rather than causal chains predominate. The unmeasured role of co-interventions such as staff education, and the wide range of how CDSS are used, reinforces the point that quantifying their undiluted effect may not be possible. In addition, there is empirical evidence to support CDSS effects being transient, highlighted by O’Connor et al (28):

> *‘once incentives (including carrot and stick) were dropped, the use of the diabetes wizard tailed off within the space of a year.’*

Caution should therefore be exercised when comparing the effect of CDSS intervention in a similar manner to medicinal products. Unlike the biological effects of medications, CDSS may behave differently even among similar populations or clinical pathways. For future AI-CDSS, an understanding of the fundamentals of human-computer interactions and why an intervention is adopted by clinicians in one context but overridden or ignored in another may be as significant a factor as the underlying performance characteristics of a CDSS, in determining the overall effect on clinical care and patient outcomes.

## CONCLUSION

In this systematic review, we have highlighted the key characteristics of existing RCTs of CDSS interventions, which notably focus on improving adherence to guidelines and take the form of active alerts. The significant variation in application of co-interventions and the underreporting of key details such as existing decision support ecosystems and metrics used to quantify alert fatigue limits the reproducibility and implementation of such findings. The similarities observed between guideline adherence trials and knowledge generation trials suggest a lack of distinction in the planning and delivery between these two groups of research studies. As digital workflow transformation continues, CDSS evidence generation and standardised implementation will be required, particularly in the era of AI-based decision support.

## Data Availability

Data extraction tables available on request.

## Author Contributions

YC & MGW conceived the study idea. TL & FA contributed to the decision-making framework. MN executed the search. DC, YC and MGW conducted the title and abstract screen. YC, MGW, JW, MD, RW, KD conducted the data extraction. YC and MGW prepared the data and wrote the first draft. All authors contributed to critical revision of the protocol and approval of the final version of the manuscript.

## Funding

This study is supported by the NIHR Central London Patient Safety Research Collaboration (CL PSRC), reference number NIHR204297. The views expressed are those of the authors and not necessarily those of the NIHR or the Department of Health and Social Care.

MGW is supported by the NIHR Biomedical Research Centre and Patient Safety Research Collaborative at UCL/H. YC is supported by the NIHR Biomedical Research Centre at UCLH. MN is supported by the UKRI CDT in AI for Healthcare, http://ai4health.io (EP/S023283/1).

## Declaration of interests

None relevant

## Notes

### Competing Interest Statement

The authors have declared no competing interest.

