## Supplemental Materials for "Design and implementation of Electronic Health Record Computerised Decision Support System (CDSS) trials: A Systematic Review"

### APPENDIX 1 – SEARCH STRATEGY

Date of search execution 9th April 2022

Database: Ovid MEDLINE(R) In-Process & Other Non-Indexed Citations, Ovid MEDLINE(R) Daily and Ovid MEDLINE(R) <1946 to Present>
Search Strategy:
--------------------------------------------------------------------------------

1  Reminder Systems/

2  reminder?.ti,ab.

3  (prompt? or alert or alerts).ti,ab.

4  or/1-3 [Reminders/Prompts]

5  (onscreen* or "on screen").ti,ab.

6  computer$.ti.

7  (computer adj2 (bedside? or bed-side? or "point of care" or screen or screens or terminal? or station? or office or desktop?)).ti,ab.
8 (information system? adj2 (bedside? or bed-side? or "point of care" or screen or screens or terminal? or station? or desktop?)).ti,ab.
9 (ipad? or i-pad? or ((tablet? or notebook?) adj2 (device? or computer?)) or netbook? or handheld?).ti,ab.
10 (mobile computer? or mobile computing or (mobile adj2 device?) or (mobile adj2 (technology or technologies))).ti,ab.

11  Computers/ or Computers, Handheld/ or Minicomputers/ or microcomputers/ or Computer Terminals/

12  or/5-11 [Onscreen/Computer]

13  4 and 12 [Set 1: Reminders & computers/screens]

14  Decision Support Systems, Clinical/ [broad, but finds relevant interventions]

15  Electronic Health Records/ and (referral? or (primary adj2 care) or general practitioner?).ti,ab,hw.

16  ((computeri?ed or web-based or electronic$) and ((decision adj2 support) or management or (patient adj2 (care or treatment?)) or (manag$ adj3 disease?))).ti.

17  ((realtime or real time) adj5 (alert? or prompt$ or notify$ or notification? or cue or cues)).ti,ab.

18  ((automat$ quer$ or software or programme or programmes or program?) adj5 (electronic adj2 (record? or medical record? or patient record? or health record?))).ti,ab.

19 (mobile computer? or mobile computing or (mobile adj2 device?) or (mobile adj2 (technology or technologies))).ti,ab.

20  ((onscreen or on-screen) adj2 reminder?).ti,ab.

21  ((patient specific or patient focussed or patient oriented or (personali?ed adj2 patient?)) and reminder?).ti. or ((patient specific or patient focussed or patient oriented or (personali?ed adj2 patient?)) adj3 reminder?).ab.

22 (reminder? and (clinical visit? or clinical encounter?)).ti. or (reminder? adj10 (clinical visit? or clinical encounter?)).ab.

23 (reminder? and ("point of care" or "point of patient")).ti. or (reminder? adj5 ("point of care" or "point of patient")).ab.

24 (reminder? and (clinical or doctor? or (during adj3 visit?) or GP or intern or interns or nurse or nurses or physician? or practitioner? or provider? or resident?)).ti.

25 (reminder? adj3 (clinical or doctor? or (during adj3 visit?) or GP or intern or interns or nurse or nurses or physician? or practitioner? or provider? Or

resident?)).ab.

26  (computer$ adj10 reminder?).ti,ab.

27  ((computer$ or electronic) adj5 (((decision$ or prescrib$ or medication$ or treatment?) adj2 support) or

reminder$ or prompt$ or cue? or alert?)).ab.

28 29 30 or 31 32 33 34 35 36 37

(display? adj10 (advice or patient-specific or recommended)).ti,ab.
(computeri?ed adj2 (suggestion? or advice or patient-specific or ((treatment or care) adj options))).ti,ab. ((prompt or prompts or prompting) and (consult$ or counsel$ or physician? or doctor? or practitioner? or nurse

nurses)).ti.
(computer$ adj2 prompt?).ti,ab.
((screen? or touch screen?) adj3 (prompt? or remind$ or warning)).ti,ab. or/14-32 [Set 2: Keywords & Proxy Concepts]
Hospital information systems/ or Clinical Pharmacy Information Systems/ Operating Room Information Systems/ or Radiology Information Systems/ Medical Order Entry Systems/
Medical Records Systems, Computerized/ or Electronic Health Records/

Page 2 of 55

38 Decision Making, Computer-Assisted/ or Diagnosis, Computer-Assisted/ or Therapy, Computer-Assisted/ or Drug Therapy, Computer-Assisted/ or Radiotherapy Planning, Computer-Assisted/
39 ((health record? or medical record? or patient chart? or treatment record?) adj2 (computeri?ed or electronic)).ti,ab.

40 ((information system? or electronic or computer system? or computer assisted or computeri?ed) adj4 (clinical or order entry or decision? or decision-making or decision support or prescribing or prescription? or drug therapy)).ti,ab.

41  Ambulatory Care Information Systems/ or Point-of-Care Systems/ or Decision Support Systems, Clinical/

42  (information system? adj3 (patient? care or "point of care" or ambulatory)).ti,ab.

43  decision support system?.ti,ab.

44  or/34-43 [Care/Decision Systems]

45  44 and 4 [Set 3: Care/Decision Systems & Reminders]

46  ((clinician? or physician? or surgeon?) adj2 discretion).ti,ab.

47  ((rapid or rapidly or immediate) adj2 (decision or choice)).ti,ab.

48  ((traditional or usual or customary) adj2 (care or dosing or prescribing or management)).ti,ab.

49  or/46-48 [Concepts related to reminder systems found in the literature]

50  44 and 49 [Set 4]

51  (randomized controlled trial or controlled clinical trial).pt. or randomized.ab. or placebo.ab. or clinical trials as

topic.sh. or randomly.ab. or trial.ti.

52  exp animals/ not humans.sh.

53  51 not 52 [Cochrane RCT Filter 6.4.d Sens/Precision Maximizing]

54  clinical trial/ or multicenter study/

55  Comparative study.pt. and ((improv$ or assessment or effectiveness or influenc$ or increase? or reduce?).ti. or

impact.ti,ab.)

56  (patient adj5 outcome?).ti,ab.

57  (or/54-56) not 52 [Filter 2]

58  ((reminder? or alerts) adj3 (electronic or clinical or on-screen or computer screen?)).ti.

59  (remind$ adj4 (physician? or nurse or nurses or counsel$ or therapist? or practitioner? or professional?)).ti.

60  (reminder? and guideline?).ti.

61  (remind$ adj4 guideline?).ab.

62  or/58-61 [Keyword Results]

63  or/13,33,45,50 [Results before filters]

64  63 and 53 [RCT Results]

65  (63 and 57) not 64 [Filter 2 Results]

66  62 not (or/52,64-65) [Keyword results]

67  remove duplicates from 64 [RCT Results]

68  remove duplicates from 65 [Filter 2 results]

69  remove duplicates from 66 [Keyword results]

70  67or68or69

71 limit 70 to 2010-current

### APPENDIX 2 – Protocol Amendments

Given the rapid growth of CDSS studies since 2019, we elected to perform a series of pragmatic deviations from our original protocol to maintain a manageable number of studies to assess. All changes were made following discussion with the authorship group and approved by the corresponding author (FA). These included: **(1)** We restricted our search to RCTs only rather than include quasi-experimental study designs. **(2)** We did not conduct any additional record searches of grey literature including technical reports, conference proceedings or working papers. **(3)** We updated the original list of 23 data variables specified in our protocol. The key differences include removal of two items (“3b. If applicable, external validation if used in multiple settings”; “11. Whether CDSS exists on an open platform”) and insertion or modification of several items (“5b. Hard stop”, “9b. CDSS target does it include patient who can also view the decision support tool?”, “9c. Is CDSS full embedded?”, “9d. Simple alert or protocol/ template”, “12a. Location of study”, “12b. Whether study is multicentre”, “14. Consent process”, “15. Unit of randomisation”, “18. Co-interventions (education & additional)”, “27. Type of outcomes measured”). The list was then arranged in a more logical manner, and grouped according to our five domains. Table S1 is the original data variable list and Table S2 is the one used in the study.

**Original list of data variables**

1. Define EHRS and version used

2. Testing e.g., for safety or working/ approach used to validate CDSS accuracy

3a. Describes results of any analysis of performance errors

3b. If applicable, external validation if used in multiple settings

4. Ongoing monitoring of CDSS performance

5. Decision applied to

6. Type of decision supported

7. CDSS target

8. Screenshots, design features e.g., font/ text

9. Active versus passive

10. Description of data quality used for CDSS including missing data

11. Whether CDSS exists on an open platform

12. Comparator

13. Table of characteristics for users of CDSS

14. Analysis of results by breakdown for CDSS users

15. Change in policy during CDSS implementation that affected CDSS workflow

16a. Evidence of elicitation of user views

16b. User training

17a. Dose of CDSS modifiable

17b. CDSS requires acknowledgement

18. Any quantified metric of CDSS fatigue (measured as outcome) or published feedback survey

19. Number of other CDSS in use at study site mentioned?

20. Co-interventions

21. Clinician ordering behaviour/ Quantify effect on clinical workflow

22. Trial protocol accessible

23. Outcome of trial

**Table S1.** Original list of data variables to be collected

| **Domain** | **Item** | **Explanation** | **Reference** |
| --- | --- | --- | --- |
| Design features and safety | 1. Define EHR and version used | Routine EHR updates can result in internal changes in variable definitions that inadvertently change definitions of data variables used by the CDSS | Finlayson SG, Subbaswamy A, Singh K, et al. The Clinician and Dataset Shift in Artificial Intelligence. N Engl J Med. 2021;385(3):283-286. doi:10.1056/NEJMc2104626 |
|  | 2. Testing e.g. for safety or working/ approach used to validate CDSS accuracy | Any mention of testing of the CDSS for issues pertaining to its intended effect or any safety testing for identification of harms | RECORD (6.1,6.2) - modified from 'code/ algorithm for patient identification, CONSORT- Routine (4a) |
|  | 3. Describe results of any analysis of performance errors. | Any reporting of CDSS performance in terms of accuracy in its deployment (e.g. displayed at the right time) | CONSORT - AI |
|  | 4. Ongoing monitoring of CDSS performance | Reporting of whether CDSS performance changed during the course of its implementation | CONSORT - Routine (Routine2) |
|  | 5a. Active versus passive | Is the CDSS visible when displayed which does not require the user to click onto it | SUNDAE (13) |
|  | 5b. Hard stop | If the user can dismiss the CDSS easily then not considered hard stop | Powers EM, Shiffman RN, Melnick ER, Hickner A, Sharifi M. Efficacy and unintended consequences of hard-stop alerts in electronic health record systems: a systematic review. J Am Med Inform Assoc. 2018;25(11):1556-1566. doi:10.1093/jamia/ocy112 |
|  | 6. Screenshots, design features e.g. font/ text | Are there details of the CDSS included sufficient to allow for replication | SUNDAE (9) |
|  | 7. Quality of the data or missing data | Any reporting of EHR data quality that would affect the performance of the CDSS | CONSORT- Routine (Routine 5), SAFER framework |
|  | 8. CDSS on any open platform? | Any detail of whether the CDSS or code for the CDSS is available publicly | SAFER framework  Pletcher MJ, Flaherman V, Najafi N, Patel S, Rushakoff RJ, Hoffman A, Robinson A, Cucina RJ, McCulloch CE, Gonzales R, Auerbach A. Randomized Controlled Trials of Electronic Health Record Interventions: Design, Conduct, and Reporting Considerations. Ann Intern Med. 2020 Jun 2;172(11 Suppl):S85-S91. |
|  | 9a. CDSS target | Detail about professional role of clinician | SUNDAE (4) |
|  | 9b. CDSS target does it include patient who can also view the decision support tool? | Collected as “is the CDSS visible to the patient” | SUNDAE (4) |
|  | 9c. Is CDSS full embedded? | EHR used for: (1) input data to CDSS for patient recognition/ trigger condition +/- (2) display of the information/ recommendation. Collected as “afferent, efferent or Both” | N/A |
|  | 9d. Simple alert or protocol/ template | Other forms of CDSS are specified if not alert-based | Jani YH, Franklin BD. Interruptive alerts: only one part of the solution for clinical decision support. BMJ Qual Saf. 2021; 30(12):933–6 |
| Context of decision | 10. Decision | Nature of clinical decision | SUNDAE (3) |
|  | 11. Type of decision supported | Guideline adherence or knowledge generation | Modified CReDECI2 (1) |
|  | 12a. Location of study | Different study sites will have different digital capability and governance frameworks to conduct trials of CDSS | Wright A, McCoy AB, Choudhry NK. Recommendations for the Conduct and Reporting of Research Involving Flexible Electronic Health Record-Based Interventions. Ann Intern Med. 2020 Jun 2;172(11 Suppl):S110-S115. |
|  | 12b. Whether study is multicentre | Was more the CDSS used at more than one study site | CONSORT - Routine |
| Study design & Implementation | 13. Comparator | Usual care, silent CDSS or CDSS+ | CReDECI2 (6) |
|  | 14. Consent process | Different models of consent were used and have been categorised. | CONSORT – Routine  Pletcher MJ, Flaherman V, Najafi N, Patel S, Rushakoff RJ, Hoffman A, Robinson A, Cucina RJ, McCulloch CE, Gonzales R, Auerbach A. Randomized Controlled Trials of Electronic Health Record Interventions: Design, Conduct, and Reporting Considerations. Ann Intern Med. 2020 Jun 2;172(11 Suppl):S85-S91. |
|  | 15a. Unit of randomisation | Cluster, patient or provider | CONSORT (3a) |
|  | 15b. Point of care randomisation | Of studies which randomised patients or providers, was randomisation conducted by, or embedded into, the EHR | Pletcher MJ, Flaherman V, Najafi N, Patel S, Rushakoff RJ, Hoffman A, Robinson A, Cucina RJ, McCulloch CE, Gonzales R, Auerbach A. Randomized Controlled Trials of Electronic Health Record Interventions: Design, Conduct, and Reporting Considerations. Ann Intern Med. 2020 Jun 2;172(11 Suppl):S85-S91. |
|  | 16. Number of other CDSS in use at study site mentioned? | Any reporting of other CDSS at the study site | CReDECI2 (11) |
|  | 17. Change in policy during CDSS implementation that affected CDSS workflow | Any reporting of external factors at the study site that could affect the CDSS workflow e.g., other change in EHR design | STARE-HI |
|  | 18a. Co-interventions (education) | Any reporting of educational interventions that explain the intended clinical purpose of the CDSS | SAFER framework |
|  | 18b. Co-interventions (additional) | Any reporting of other interventions that support the CDSS, including local leadership | STARE-HI, StARI |
| Relevant human factors | 19. Table of characteristics for users of CDSS | Difference between professional groups/ hierarchy? | StARI |
|  | 20. Analysis of results by breakdown for CDSS users |  | SAFER framework |
|  | 21a. Evidence of elicitation of user views | Any reporting of user feedback on CDSS design before the start of the study | HOT-fit framework |
|  | 21b. User training | Any reporting of staff education on how to use the CDSS, which occurred before the start of the study | HOT-fit framework |
|  | 22. Was alert dose modifiable | Only applicable to subset of CDSS which were alerts: any option for users to alter the sensitivity of the CDSS being triggered | Kawamoto K, McDonald CJ. Designing, Conducting, and Reporting Clinical Decision Support Studies: Recommendations and Call to Action. Ann Intern Med. 2020 Jun 2;172(11 Suppl):S101-S109. |
|  | 23. Clinician ordering behaviour | Any reporting of metrics which monitor CDSS effect on clinicians e.g., % alerts dismissed or referred to another provider, Log analysis and time spent on EHR, or eye tracking etc | SAFER framework |
|  | 24. Alert fatigue | Only applicable to subset of CDSS which were alerts: any reporting of metrics related to alert fatigue such as user feedback survey or quantitative measures e.g., high dismissal rate | Kawamoto K, McDonald CJ. Designing, Conducting, and Reporting Clinical Decision Support Studies: Recommendations and Call to Action. Ann Intern Med. 2020 Jun 2;172(11 Suppl):S101-S109.  McGreevey JD 3rd, Mallozzi CP, Perkins RM, Shelov E, Schreiber R. Reducing Alert Burden in Electronic Health Records: State of the Art Recommendations from Four Health Systems. Appl Clin Inform. 2020 Jan;11(1):1-12. |
| Study outcomes and reporting | 25. Trial protocol accessible | Was the study protocol available | SPIRIT & CONSORT |
|  | 26. Outcome | For the primary outcome, was the study reported as positive or negative | CONSORT |
|  | 27. Type of outcomes measured | Clinical or process measure | CONSORT Routine (6a) |

**Table S2.** Updated list of data variables to be collected. The explanation column was used to guide the authors allocated to data extraction for eligible studies. Reference to established reporting guidelines or recommendations are made where relevant

### APPENDIX 3 –Elaboration of scoring

The following examples are given to elaborate on how certain variables from the data extraction form were adjudicated. This includes those that were most open to interpretation, and were agreed by the study group.

5a & 5b. Definition of Active alert/ hard stop

There were some key examples from the list of studies which acted as precedent for the definition of active alert. For example:

*Terrell KM, Perkins AJ, Hui SL, Callahan CM, Dexter PR, Miller DK. Computerized decision support for medication dosing in renal insufficiency: a randomized, controlled trial. Ann Emerg Med 2010;56:623-629.*

The alert was active but this was determined by the screenshot included rather than an author description.

*Andruchow JE, Grigat D, McRae AD, Innes G, Vatanpour S, Wang D, et al. Decision support for computed tomography in the emergency department: a multicenter cluster-randomized controlled trial. CJEM, Can j emerg med care. England; 2021; 23(5):631–40.*

The CDSS defined as an alert was active based on the description that it ‘opened in an external window’. However, however it did not require acknowledgement and therefore was deemed ‘non-hard stop’. 91 of 96 Alert-based CDSS were pure alerts, 5 were mixed alert and templates.

1. Type of Decision

The distinction between whether a CDSS supported guideline adherence or was knowledge generating was defined according to the following boundary:

1. If the CDSS was aiming to change clinician behaviour toward taking an action that was proven to be best practice, this was GDT. For example, an alert to prescribe prognostic medication in patients with confirmed heart failure
2. If the CDSS was aiming to investigate whether earlier intervention of known beneficial, goal directed therapy improved patient outcomes For example, an alert to highlight the presence of an acute kidney injury and to recommend the clinical team to act accordingly, this was knowledge generating.

In the second example, the hypothesis of earlier intervention leading to better outcomes did not have a robust answer prior to the conduct of the trial and was therefore knowledge generating.

21a. Definition of Evidence of Elicitation of views

In terms of evidence to support fulfilment of whether the views of users was incorporated, the gold standard was by Gill et al:

Gill JM, Mainous AG, 3rd, Koopman RJ, et al. Impact of EHR-based clinical decision support on adherence to guidelines for patients on NSAIDs: a randomized controlled trial. Ann Fam Med 2011;9:22-30.

“The EHR-based clinical decision support form was pilot tested with focus groups of clinicians who used the same EHR but were not study participants. Their feedback was used to modify the form and create the final version.”

However, the requirement for feedback to be reported to be used to modify the design of the CDSS was not needed.

27. Definition of types of outcome measured

For whether the study incorporated a clinical or process outcome as their primary outcome, this was clear in the majority of the reports. For some of the CDSS that aimed to improve prescribing behaviour, for example antibiotic stewardship, a process outcome was defined as observing a decrease in the rate of antibiotics while a clinical outcome was defined as the rate of infection.
